# Assessment of trends and determinants of under-five mortality among children born to older women: Evidence from Ethiopian Demographic and Health Surveys

**DOI:** 10.1101/2025.02.05.25321717

**Authors:** Tamerat Denekew Temesegen, Tariku Dejene Demissie, Solomon Abrha Damtew

## Abstract

**Introduction:** Mortality has long been used as an indicator of the level of socio-economic development of a country. Global attention has been drawn to under-five mortality through the Sustainable Development Goals (SDGs). The burden of under-five mortality of children is still not fairly divided. Two regions account for around 80% of under-five deaths among children: sub-Saharan Africa and South Asia. Ethiopia is one of six nations that account for half of the world’s under-five mortality.

**Methods:** The Ethiopia Demographic and Health Survey (EDHS) from the years 2000, 2005, 2011, and 2016 provided nationally representative cross-sectional data. Six thousand one hundred nighty nine (6,199) children born to older women within the five years prior to the study formed the data. Home interviews were used to gather demographic data, such as mother and child characteristics, socioeconomic factors, and environmental variables. Frequencies were computed to characterize the study participants Multiple-level binary regression analysis was utilized to get the Adjusted Odds Ratio (AOR) and its 95% confidence interval (CI).

**Result:** The absolute number of under five deaths is 128/1000 live births in the year 2000 which reduce to 56/1000 live births in 2016. This study identified factors contributing for under-five mortality among children of women aged 35 to 49 years old. Sex of the child (female_ 0.65 (0.53, 0.79)), late age at first birth, 31 to 4o years at first birth 0.34 (0.11, 0.98), family size, having a family size of 6 to 10 0.18 (0.14, 0.24), longer birth interval, greater than 3 years 0.35 (0.26, 0.47) and ANC visits 1 to 3 ANC vests 0.65 (0.5, 0.84) lowered the odds of under-five mortality among children of relatively elders. On the other hand, Women aged 35 to 49 years who gave a twin Birth 6.15 (3.94, 9.6)), those with number of births in the last five years having 4 children (2.64 (1.15, 6.06)), those with number of children ever born, having 9 or above children 3.79 (2.1, 6.84), those who gave birth at late age the index child, mother gave birth of the index child at late age (45 to 49) 2.13 (1.55, 2.93) had higher odds for their children experiencing child death before they celebrated their fifth birthday.

**Conclusion:** The absolute number of under five deaths of older women is 128/1000 live births in the year 2000 which reduce to 56/1000 live births in 2016.Female children, children born from first from elder mothers, those children residing am family with large size, children born with longer birth interval and children for whom their mothers received ANC visits were found to Have lower odds of mortality.

## Background

The World Health Organization defined under five mortality as the mortality rate of children aged under 5 years is the probability that a child born in a specific year or period will die before reaching the age of 5 years, subject to the age-specific mortality rates of that period.

Under five mortality was set as one of the country’s heath system infrastructure indicator in terms of health equity and health system challenges along with economic growth and development build on the success of MGD (1, 2), SDG goal 3 target 3.2 setted to end preventable deaths of newborns and children under 5 years of age, with all countries aiming to reduce under-5 mortality to at least as low as 25 per 1,000 live births by 2030 (3, 4).

Ethiopian is one of the East African countries who have shown remarkable progress in achieving the MDG goals notable the promising reduction in undefined mortality, The under-five mortality rate in Ethiopia was 67 deaths per 1000 live births, according to the Ethiopian Demographic and Health Survey 2016 this means that one out of every fifteen Ethiopian children dies before reaching the age of five. Under-five mortality in the country fell from 166 deaths per 1000 live births in 2000 to 67 deaths per 1000 live births in 2016, a 60% decrease in under-five mortality over 16 years (5).

Child mortality rates in Ethiopia have declined over time, the progress has not been as rapid as hoped. According to the 2016 Ethiopian Demographic and Health Survey (EDHS), the under-five mortality rate for the 5-year period preceding the survey was 67 deaths per 1,000 live births. This means that approximately 1 out of every 15 children in Ethiopia does not survive to their fifth birthday (6–9). This is one of the difficult issues the nation needs to solve because the rate is still high.

Knowing the socioeconomic factors that contribute to under-five mortality in Ethiopia can help identify the segments of the population that require assistance in order to lower death rates more rapidly. The rate of decrease in under-five mortality is still rather high, and more work is needed to eliminate obstacles to under-five survival, even though several research have been carried out to identify variables related with under-five mortality in Ethiopia.

Therefore, the Federal democratic republic of Ethiopia Health Mister have designed and implementing various policy strategies to reduce under five mortality as clearly speculated in the countries Newborn care guidelines and in the integrated management of Common childhood illness guideline (10–12). This is reflected in the countrýs recent health sector transformation plan as well (13). Moreover, Improvement in health preventive and curative service provision, notably free of charge under five children treatment at health post level. Access to modern health services for the mother during pregnancy, childbirth and postnatal period have contributed its share in the reduction of under-five mortality. The government show further commitment for newborn care including NICU and teaching hospitals train midlevel professionals such as neonatal nurse and resident programs fill in the gap in skilled man power (14, 15). Access to health Insurance is another commitment for as health care equity and availing health for all citizens’ (16–18).

There are studies conducted using EDHS on the factors and magnitude of under-five mortality (19–22), however, the aim of this study was to quantify under five mortality among children of mother who are aged 35 to 49 and who have their child by then. In addition, this study also tried to address unadjusted trend of under-five mortality over the last 16 years. Hence, this study used EDHS data 2000 to 2016 to measure the level of under-five mortality to assess the overall trend and identify factors contributing for this variation. The finding could be vital for the MOH and key actors in their collaborative endeavor to track the achievement of SDG 3.2 (4).

## Methods

### Study design and Data Sources

The data for this study have been obtained from four consecutive Ethiopia Demography and Health Surveys (EDHSs) conducted in 2000, 2005, 2011 and 2016. The 2000, 2005, 2011 and 2016 Ethiopia Demographic and Health Survey (EDHS) were implemented by the Central Statistical Agency (7–9). By virtue of its mandate, the CSA had conducted the surveys in collaboration with the Federal Ministry of Health (FMoH) and the Ethiopian Public Health Institute (EPHI) with technical assistance from ICF international, and financial as well as technical support from development partners. All actors in this effort have exerted themselves to get reliable, accurate, and up-to-date data to measure the success of the national development agenda-growth and transformation plan II as well as the sustainable development goals. The 2016 survey was conducted from January 18, 2016, to June 27, 2016, based on a nationally representative sample that provides estimates at the national and regional levels and for urban and rural areas (9).

The 2000, 2005, 2011 and 2016 Ethiopia Demographic and Health Surveys, were designed to provide estimates for the health and demographic variables of interest for the following domains: Ethiopia as a whole; urban and rural areas (each as a separate domain); and 11 geographic administrative regions (nine regions namely: Tigray, Affar, Amhara, Oromiya, Somali, Benishangul-Gumuz, Southern Nations, Nationalities and Peoples (SNNP), Gambela and Harari regional states and two city administrations :Addis Ababa and Dire Dawa) (9).

### Study population and sample size

The sampling frame used for the 2000, 2005, 2011 and 2016 EDHS data is the Ethiopia Population and Housing Census (EPHC) conducted in 1994 (for the first two surveys) and in 2007 (for the latter two surveys) by the Central Statistical Agency (5, 7–9). The 2016 EDHS sample was stratified and selected in two stages. Each region was stratified into urban and rural areas, yielding 21 sampling strata. Samples of EAs were selected independently in each stratum in two stages. In the first stage, a total of 645 EAs (202 in urban areas and 443 in rural areas) were selected with probability proportional to EA size (based on the 2007 EPHC) and with independent selection in each sampling stratum. In the second stage of selection, a fixed number of 28 households per cluster were selected with an equal probability systematic selection from the newly created household listing. A total of 18,008 households were selected for the sample, of which 17,067 were occupied. Of the occupied households, 16,650 were successfully interviewed, yielding a response rate of 98% (5, 9).

In the interviewed 16,583 households, eligible women aged 15-49 were identified for individual interview; complete interviews were 15,683, conducted for yielding a response rate of 95 percent, 4,239 women were older women (aged 35 to 49) out of this 1,738 women who gave birth in the five years preceding the survey were included in this analysis. Information for this study were take from the birth history section of the Women’s Questionnaire (5).

In this study, the 2000, 2005, 2011 and 2016 EDHS data are used to describe the trend of under-five mortality in Ethiopia and to analyze determinants and variation of under-five mortality of older women by background characteristics.

### Sample Selection and sampling procedure

#### Sampling frame

The population enumeration areas (EAs) of the 1994 census was used as a sampling frame for drawing the sample for the 2000 and 2005 EDHS, while the 2007 Population and Housing census conducted by the CSA was used as a sampling frame from which the 2011 and 2016 EDHS sample was drawn (5).

#### Sampling technique and procedure

In the census frame, each of the 11 administrative areas is subdivided into zones and each zone into weredas which then sub-divided into kebels which are then further sub divided into convenient areas called census enumeration areas (EAs).Selected population census enumeration areas (EA) which were used as primary sampling unit and selected households were secondary sampling units whereas household members were observational or study units (5).

A complete household listing operation was carried out in all the selected EAs to provide a sampling frame for the second-stage selection of households. To avoid an uneven sample allocation among regions, the sample was allocated by region in proportion to the square root of the region’s population size instead of proportional sample allocation since this procedure yielded a distribution in which 80 percent of the sample came from three regions alone (5).

#### Data Collection

The data sets used for this study was obtained from measure DHS project (www.dhsprogram.com). Data were collected by visiting households and conducting face-to-face interviews to obtain information on demographic characteristics, wealth, nutritional and sexual behavior, among other data, in the respective data collection time for the fourth EDHS surveys (5, 7).

### Variables

#### Dependent (Outcome) Variable

The Dependent (Outcome) variable for this study is under-five child mortality of older women. Under-five mortality is defined as the probability of dying before completing the fifth birthday. Thus, the outcome variable is the child event before reaching five years of age, which is dichotomous and coded as 1 if the child died in the five years before the survey and 0 if alive.

➢ Under-five death (0=Yes, 1= No)

### Independent Variables

The explanatory variables included in this study are based on the Mosley and Chen (1984) determinants of childhood morbidity and mortality framework for developing countries, experiences from the available similar studies reviewed above and available data on the subject.

The main predictor variables of under-five child mortality explored include **Demographic, Socioeconomic and Environmental factors**.

#### The demographic variables/factors include:-

➢ Sex of child (1=male and 2= female)
➢ Age of child in month
➢ Type of birth (0=Single birth and 1=Multiple birth)
➢ Birth order number (1= first birth, 2= between 2-4, 3= above 5)
➢ Preceding birth interval in month (0= First birth, 1=below 24 months, 2=between 24-35 months and 3= above 36 months)
➢ Age of mother at first birth (1=under 20 years, 2= between 21-30 years and 3= 31-40 years, 4= above 41 years)
➢ Family size (Number of HH members) (1= between 1-5, 2=between 6-10 and 3= above 11)
➢ Breastfeeding status (1=ever breastfeed and 2= never breast feed).

#### The socioeconomic variables/factors include:-

➢ Mothers’ education level (0=no education, 1=primary education, 2=secondary education and above)
➢ Wealth index (economic status of HH) (1=Poor, 2=Medium and 3=Rich)
➢ Region(1=Tigray,2=Afar,3=Amhara,4=Oromia,5=Somali,6=Benshangule-Gumuz,7=SNNP, 8=Gambella,9=Harari,10=Addis Ababa and 11=Diredawa)
➢ Place of residence (1=Urban and 2=Rural).

#### And the variables that are classified as environmental variables include:-

➢ Availability of toilet facility (0=improved facility and 1=unimproved facility and 2=no toilet facility).
➢ Source of drinking water (0=protected source and 1=unprotected source)
➢ Place of delivery (0=at home and 1=at health center)

### Statistical Analysis

After data were checked for inconsistencies, missing values and outliers, analysis were performed using STATA version 16.0 statistical software. Variables were computed and recoded to fit the objective of this study.

Multilevel binary logistics were used to analyze the data. First the individual women sample weight were calculated by dividing v005 by 1,000,000. Then plan analysis were prepare by putting the strata, primary sampling units and sample weight for each data set since the complex sample analysis requires preparation of plan for analysis for the data set before conducting the analysis. The complex sample commands in STATA were used to declare the strata, primary sampling unit and sample weight. This were done so that STATA were not assume that the data came from simple random sampling, ignores the stratification and clustering which increases the significance and decreases the standard error of point estimates. It is worth noting that the point estimates are similar whether complex sample analysis is used or not in stratified cluster sampling.

Absolute counts, percentages and odds ratios present in this study were all weighted for the sampling probabilities.

Frequencies and proportions were computed for description of the study population in relation to selected socio-demographic variables using complex sample frequency and descriptive analysis. The results were present in the form of tables and figures.

To see the association of U5MR and selected socio-demographic variables [age, wealth index, marital and educational status, parity, region of residence and occupation], complex sample bivariate and multivariate logistic regression model were used. Results were present in the form of odds ratios (OR) with 95% confidence intervals.

### Data Quality Control and Management

After accessing the data sets the data were explore to examine missing values and outliers. Data completeness were be checked by running frequency. The data quality control and management employed in the surveys is well documented in the EDHS full reports. Briefly, data were collected by visiting households and conducting face-to-face interviews using pretested questionnaire after intense training has been given for datacollectors.

### Ethical Considerations

The study was conducted after obtaining ethical clearance from Addis Ababa University, and consent from Measure DHS project. As this manuscript is based on secondary data, the letter of permission to access the data sets was obtained from measure DHS project (ORC Macro) include the like that allows you to download the data sets .

## Result

### Demographic Distribution of women

Among women in the age category 40 to 44 years 46.7%, 45.1%, 47.4% and 44.3% of the samples accounted for the year 2000, 2005, 2011 and 2016 respectively. Likewise, among those households headed by women close to 1 in 7 household (14.8%,16.7% and 15.5%) were female in gender for the 2000, 2011 and 2016 year surveys respectively were headed by women while one in 10 (12.8%) of households were women headed for the 2005 year. In a similar fashion, the proportion of women with family size up to 5 of less was 17.2%, 11.8%, 12.7 and 14.8% for the respective survey years among sample of women in this category among those reported a family size of 5 or less.

Regarding the number of ever born women reported that four in ten women; 40.9% for 2000 survey year, 40.2 for 2005 43.0 for 2000 reported that their number of live births is 7 or less while one in 2 women reported same number in the year 2016. The maximum number of births in last 5 years was reported 4 in all survey years, accordingly 44.5%,43.3%,46.1% and 40.1% of women reported that they had a couple of births in the last 5 years for the respective survey years. Concerning age at first birth, 6 in 10 women reported that they gave birth at age 20 years or before in the later 3 survey, while 69.4% for the 2000 year survey.

**Table 1.**
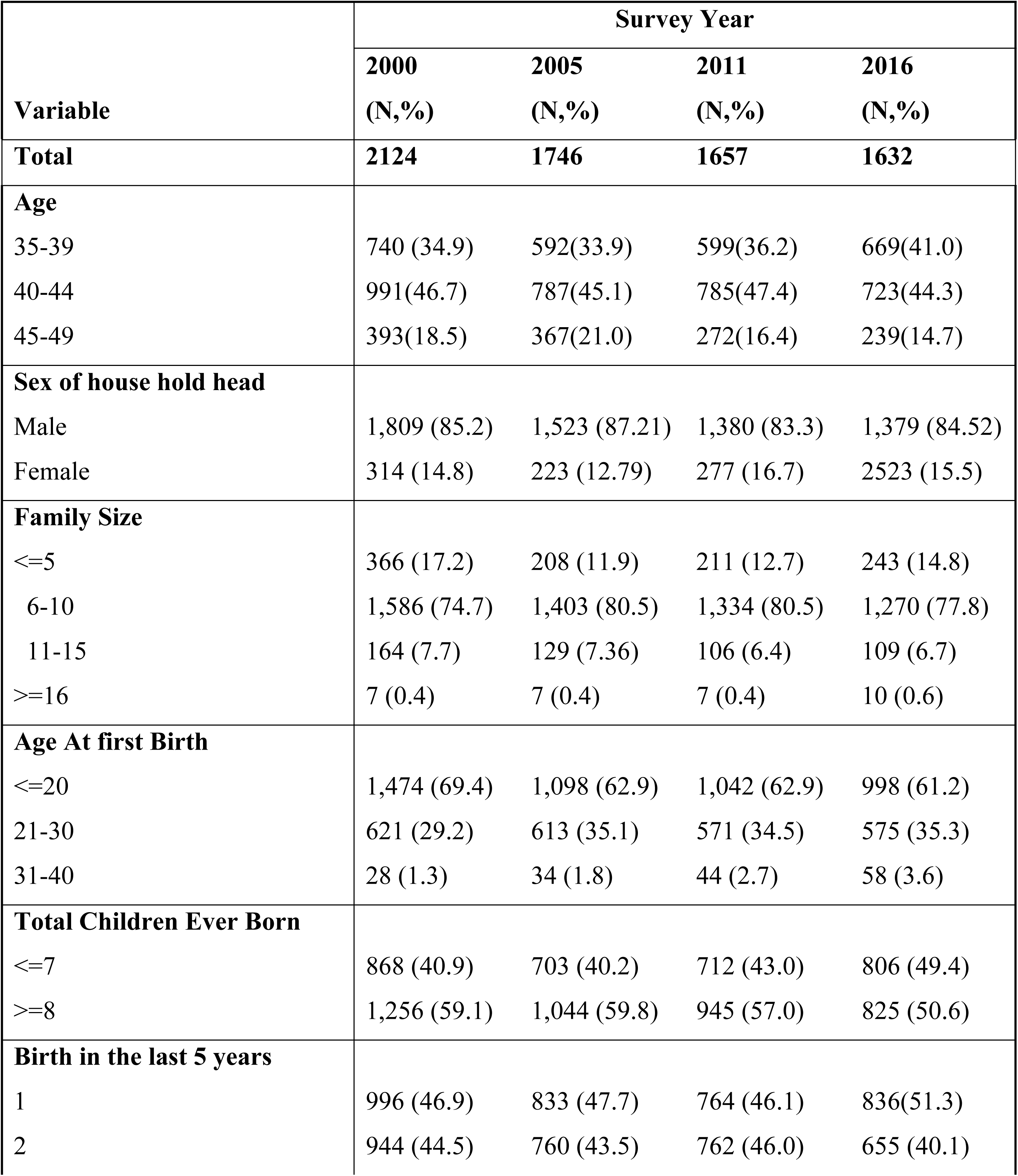
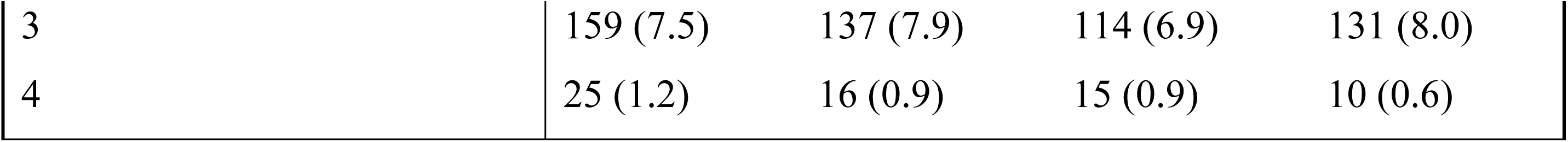
Demographic Distribution of the Women aged 35 to 49 at the birth of index child.

### Demographic Distribution of Child

Among newborns, one in two new born were female in gender for the respective years (46.5%, 51.8%, 47.2% and 50.2%). Similarly, less than one in 25 new born were twins in the respective surreys. Among those new borns who died before celebrating their first birth day more than ¾ (79.6%, 82.6%, 85.9% and 87.5%) of newborns contributed from each survey years. Likewise, nearly one third 32.5% in 2000, 29.0% in 2005, 28.7% in 2011 and one in 4 (25.9%) in 2016 were born 24 to 46 months after the preceding child.

**Table 2.** Demographic Characteristics of Index Child.

| Variable | Survey Year |  |  |  |
| --- | --- | --- | --- | --- |
|  | 2000<br>(N,%) | 2005<br>(N,%) | 2011<br>(N,%) | 2016<br>(N,%) |
| **Total** | **2124** | **1746** | **1657** | **1632** |
| **Sex of index Child** |  |  |  |  |
| Male | 1,137 (53.5) | 842 (48.2) | 875 (52.8) | 813 (49.8) |
| Female | 987 (46.5) | 905 (51.8) | 781 (47.2) | 819 (50.2) |
| **Types of Birth** |  |  |  |  |
| Single | 2,072 (97.6) | 1,679 (96.2) | 1,619 (97.7) | 1,568 (96.1) |
| Twin | 52 (2.4) | 67 (3.8) | 38 (2.3) | 63 (3.9) |
| **Birth Order of Index Child** |  |  |  |  |
| 1st birth | 6 (0.3) | 6 (0.3) | 15 (0.9) | 8 (0.5) |
| 2nd birth | 12 (0.6) | 17 (0.9) | 32 (1.9) | 25 (1.5) |
| 3-4 births | 115 (5.4) | 103 (5.9) | 99 (6.0) | 163 (10.0) |
| 5 and above Births | 1,991 (93.8) | 1,621 (92.8) | 1,511 (91.2) | 1,436 (88.0) |
| **Age of index Child(Years)** |  |  |  |  |
| 0 Years | 371 (20.0) | 389 (24.6) | 412 (26.7) | 325 (21.1) |
| 1 Years | 343 (18.5) | 289 (18.3) | 247 (16.0) | 341 (22.2) |
| 2 Years | 336 (18.1) | 274 (17.3) | 287 (18.7) | 290 (18.8) |
| 3 Years | 418 (22.6) | 310 (19.6) | 301 (19.5) | 283 (18.4) |
| 4 Years | 385 (20.8) | 319 (20.2) | 293 (19.0) | 301 (19.5) |
| **Age at Death** |  |  |  |  |
| Within the first 12 Months | 216 (79.6) | 136 (82.7) | 100 (85.9) | 80 (87.5) |
| 13 - 24 Months | 37 (13.6) | 17 (10.1) | 13 (11.3) | 5 (5.5) |
| After 25 Months | 18 (6.8) | 12 (7.3) | 3 (2.8) | 6 (7.0) |
| **Preceding Birth Interval** |  |  |  |  |
| Up to 23 Months | 274 (12.9) | 225 (13.0) | 212 (12.9) | 254 (15.6) |
| 24-36 Months | 689 (32.5) | 504 (29.0) | 471 (28.7) | 421 (25.9) |
| More than 37 Months | 1,156 (54.6) | 1,011 (58.1) | 958 (58.4) | 949 (58.5) |

### Socio Economic

Among women with no education 95% were from 2000 year survey while 90.5%, 79.4% and 82.6% were from the later surveys respectively. Concerning marital status among those widowed, divorced and no longer living together categories less than 1 in 25 women accounted for each year of survey respectively. Of those residents of highland regions More than 90% (96.1%, 93.0%, 93.2% and92.0%) of respondents were resident these areas of the country for the respective survey years. Among Orthodox religion followers 1 in 4 (46.5%, 41.8% and 41.1%) of women were from the later three survey while 520. % was from 2000 years sample. More than 90% of the sample comes from the rural clusters of the country.

**Table 3.** Socio Economic Distribution of the Mothers and Index Child by Survey Year.

| Variable | Survey Year |  |  |  |
| --- | --- | --- | --- | --- |
|  | 2000<br>(N,%) | 2005<br>(N,%) | 2011<br>(N,%) | 2016<br>(N,%) |
| Total | 2124 | 1746 | 1657 | 1632 |
| **Highest Education Level** |  |  |  |  |
| No education | 2,017 (95.0) | 1,580 (90.5) | 1,315 (79.4) | 1,346 (82.5) |
| Primary | 82 (3.9) | 132 (7.6) | 320 (19.3) | 240 (14.5) |
| Secondary | 21 (1.0) | 28 (1.6) | 13 (0.8) | 27 (1.7) |
| Higher | 4 (0.2) | 6 (0.3) | 9 (0.5) | 17 (1.04) |
| **Marital Status** |  |  |  |  |
| Never in union | 2 (0.1) | 4 (0.2) | 1 (0.04) | 2 (0.2) |
| Married | 1,946 (91.6) | 1,625 (93) | 1,448 (87.4) | 1,544 (94.6) |
| Living with partner | 25 (1.2) | 20 (1.2) | 78 (4.7) | 12 (.8) |
| Widowed | 62 (2.9) | 53 (3.0) | 67 (4.1) | 35 (2.2) |
| Divorced | 56 (2.6) | 22 (1.3) | 46 (2.8) | 27 (1.7) |
| No longer living together | 32 (1.5) | 21 (1.2) | 15 (0.9) | 12 (0.7) |
| **Region** |  |  |  |  |
| Urban | 26 (1.2) | 22 (1.3) | 18 (1.1) | 33 (2.1) |
| Mainly Agrarian | 2,042 (96.1) | 1,624 (93) | 1,544 (93.2) | 1,501 (92) |
| Mainly Pastoralist | 56 (2.6) | 101 (5.8) | 94 (5.7) | 97 (6) |
| **Place of Residence** |  |  |  |  |
| Urban | 145 (6.8) | 88 (5.0) | 175 (10.6) | 128 (7.9) |
| Rural | 1,979 (93.2) | 1,658 (95.0) | 1,482 (89.4) | 1,503 (92.1) |
| **Religion** |  |  |  |  |
| Orthodox | 1,105 (52.0) | 813 (46.5) | 693 (41.8) | 6,701 (41.1) |
| Catholic | 13 (0.6) | 30 (1.7) | 17 (1.1) | 25 (1.5) |
| Protestant | 278 (13.1) | 328 (18.7) | 332 (20.0) | 334 (20.4) |
| Muslim | 647 (30.5) | 530 (30.4) | 545 (32.9) | 567 (34.7) |
| Other | 80 (3.8) | 46 (2.7) | 70 (4.3) | 36 (2.2) |

### Environmental and Health Related

Among the total who delivered at heath facility, 3.4% were from the 2000 sample, while 3.5%, 6.4% and 21.3% were from the 2005, 2011 and 2016 samples. Form those who started ANC follow up 3 month or less after conception one in 4 women (24.8% and 23.6%) were from the 2000 and 2011 sample while one in 6 (16.1% were from the 2005 and 3 in 10 (31.5%) of women were from the lasted EDHS survey. Regarding their FP use among those who reported who used contraceptive 6.7%, 11.2%, 18.2% and 27.7% were form the respective samples.

**Table 4.**
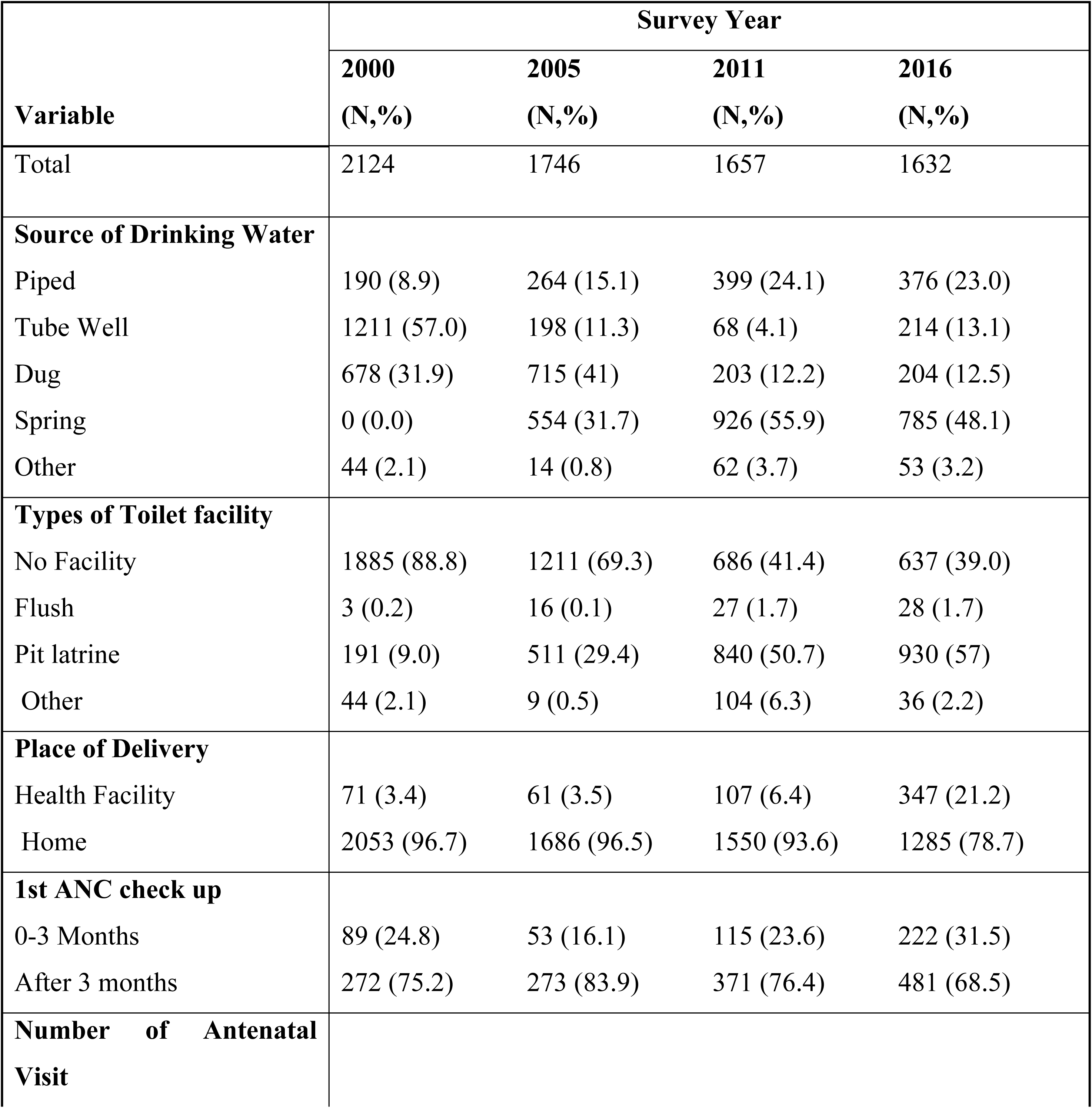
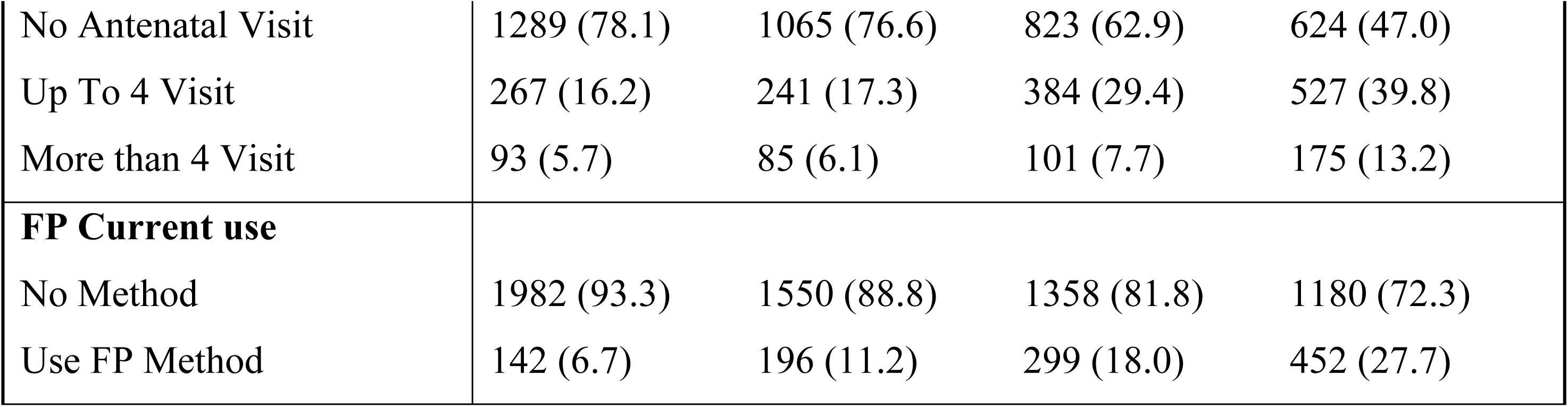
Environmental and Health Related Distribution of the Women aged 35 to 49 and child pars by Independent variables.

| **Variable** | **Survey Year** |  |  |  |
| --- | --- | --- | --- | --- |
|  | **2000**<br>**(N,%)** | **2005**<br>**(N,%)** | **2011**<br>**(N,%)** | **2016**<br>**(N,%)** |
| Total | 2124 | 1746 | 1657 | 1632 |
| **Source of Drinking Water** |  |  |  |  |
| Piped | 190 (8.9) | 264 (15.1) | 399 (24.1) | 376 (23.0) |
| Tube Well | 1211 (57.0) | 198 (11.3) | 68 (4.1) | 214 (13.1) |
| Dug | 678 (31.9) | 715 (41) | 203 (12.2) | 204 (12.5) |
| Spring | 0 (0.0) | 554 (31.7) | 926 (55.9) | 785 (48.1) |
| Other | 44 (2.1) | 14 (0.8) | 62 (3.7) | 53 (3.2) |
| **Types of Toilet facility** |  |  |  |  |
| No Facility | 1885 (88.8) | 1211 (69.3) | 686 (41.4) | 637 (39.0) |
| Flush | 3 (0.2) | 16 (0.1) | 27 (1.7) | 28 (1.7) |
| Pit latrine | 191 (9.0) | 511 (29.4) | 840 (50.7) | 930 (57) |
| Other | 44 (2.1) | 9 (0.5) | 104 (6.3) | 36 (2.2) |
| **Place of Delivery** |  |  |  |  |
| Health Facility | 71 (3.4) | 61 (3.5) | 107 (6.4) | 347 (21.2) |
| Home | 2053 (96.7) | 1686 (96.5) | 1550 (93.6) | 1285 (78.7) |
| **1st ANC check up** |  |  |  |  |
| 0-3 Months | 89 (24.8) | 53 (16.1) | 115 (23.6) | 222 (31.5) |
| After 3 months | 272 (75.2) | 273 (83.9) | 371 (76.4) | 481 (68.5) |
| **Number of Antenatal Visit** |  |  |  |  |
| No Antenatal Visit | 1289 (78.1) | 1065 (76.6) | 823 (62.9) | 624 (47.0) |
| Up To 4 Visit | 267 (16.2) | 241 (17.3) | 384 (29.4) | 527 (39.8) |
| More than 4 Visit | 93 (5.7) | 85 (6.1) | 101 (7.7) | 175 (13.2) |
| **FP Current use** |  |  |  |  |
| No Method | 1982 (93.3) | 1550 (88.8) | 1358 (81.8) | 1180 (72.3) |
| Use FP Method | 142 (6.7) | 196 (11.2) | 299 (18.0) | 452 (27.7) |

### Trend

The overall under five mortality trend has shown a decreasing trend over the 16 years period from 127/1000 live births in 2000 to 56 /1000 live births in 2016 which is the reflection for the decrement in infant mortality form 91/1000 live births to 44/1000 live births over the same period which in turn is likely to be resulted from the corresponding decrease in neonatal mortality from 51/1000 live births to 36/1000 live births (Fig 1). The mainly agrarian (Amhara, Oromia, Tigiray and SNNPR) under five mortality trend over the last one and half decade is more or less similar with the national trend at least in absolute terms . On the contrary the mainly urban comprising the three metropolis namely: Addis Ababa, Harar and Dire Dawa was found a bit higher than the national average under five mortality pattern while the mainly Pastoralists :-(Afar, Benshangul Gumuz, Gambella and Somali) under five mortality showed a lower trend from both over the 16 years period at least in terms of absolute count (Fig 1).

**Fig 1:**
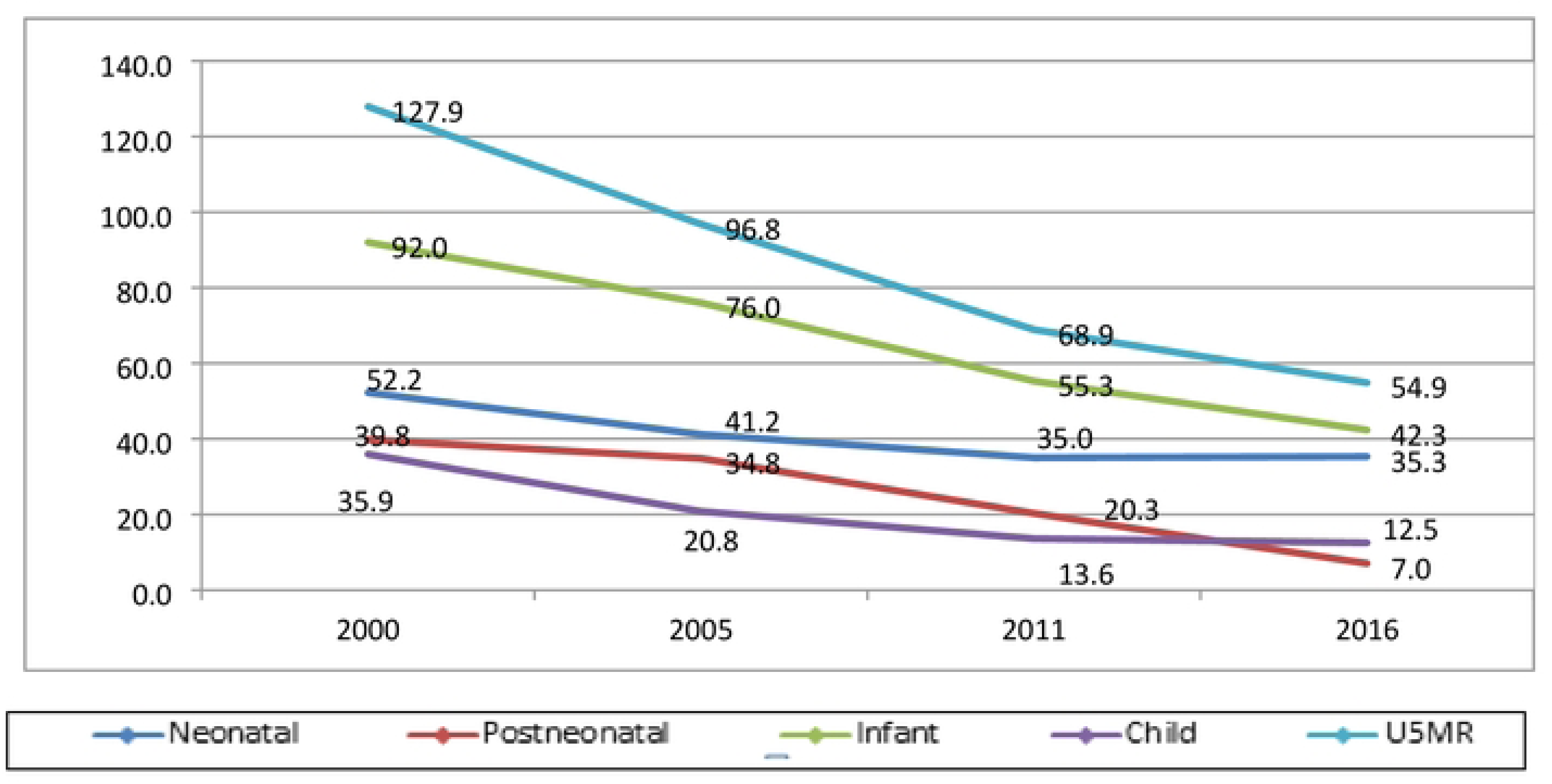
Trend of under-five, neonatal, post neonatal, infant and child mortality among children born to older women in national level: Evidence from Ethiopian Demographic and Health Surveys.

The overall mainly agrarian region under five mortality trend and desegregated trends are more or less similar with the respective national figures ((Fig 2).

**Fig 2:**
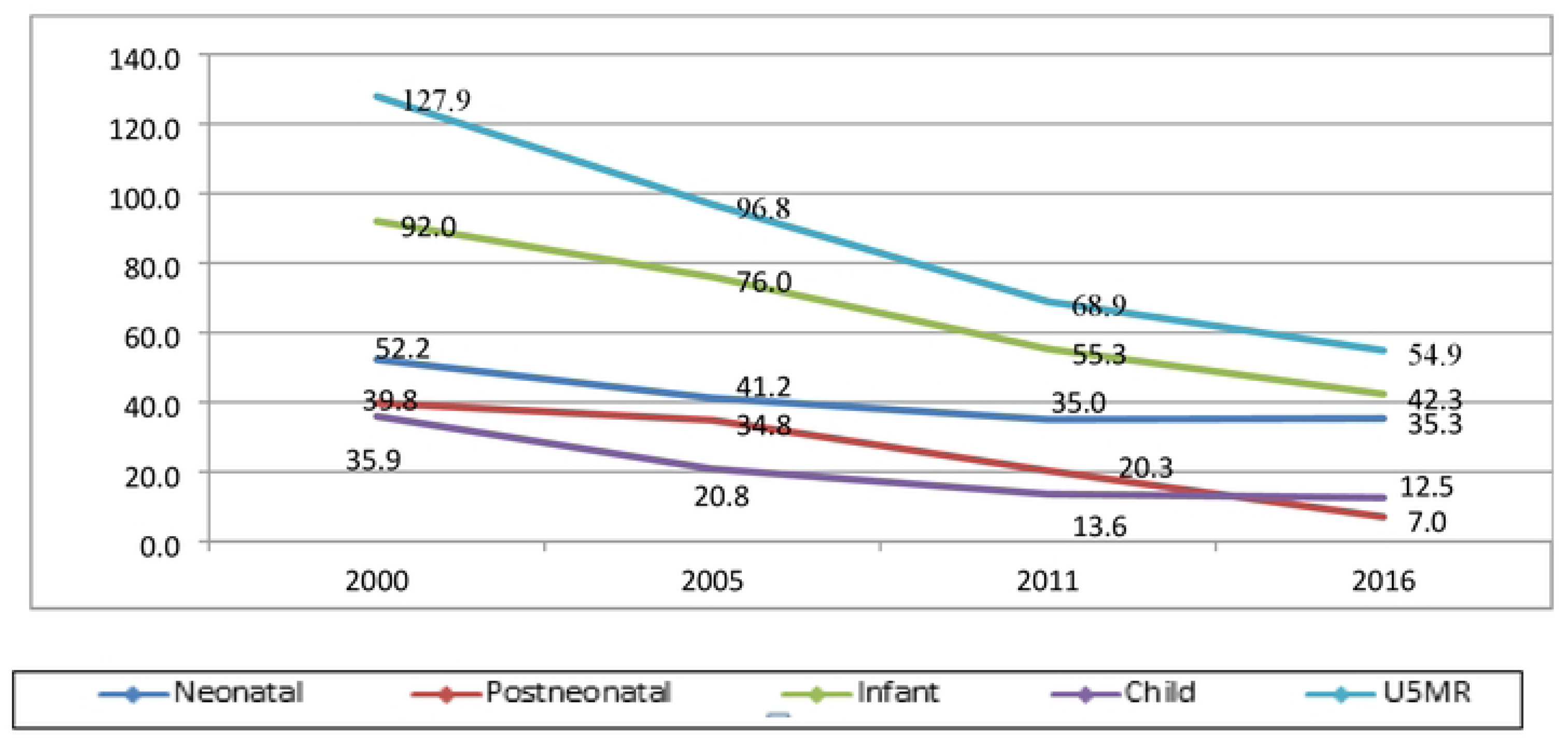
Trend of under-five, neonatal, post neonatal, infant and child mortality among children born to older women in Mainly Agrarian Regions: Evidence from Ethiopian Demographic and Health Surveys.

The overall urban under five mortality trend has shown a decreasing trend over the last 16 years from 62/1000 live births to 13 /1000 live births which is the reflection for the decrement in neonatal mortality form 52/1000 live births to 35/1000 live births over the same period resulted from the decrease in infant mortality 144/1000 live births to 14/1000 live births (Fig 3).

**Fig 3:**
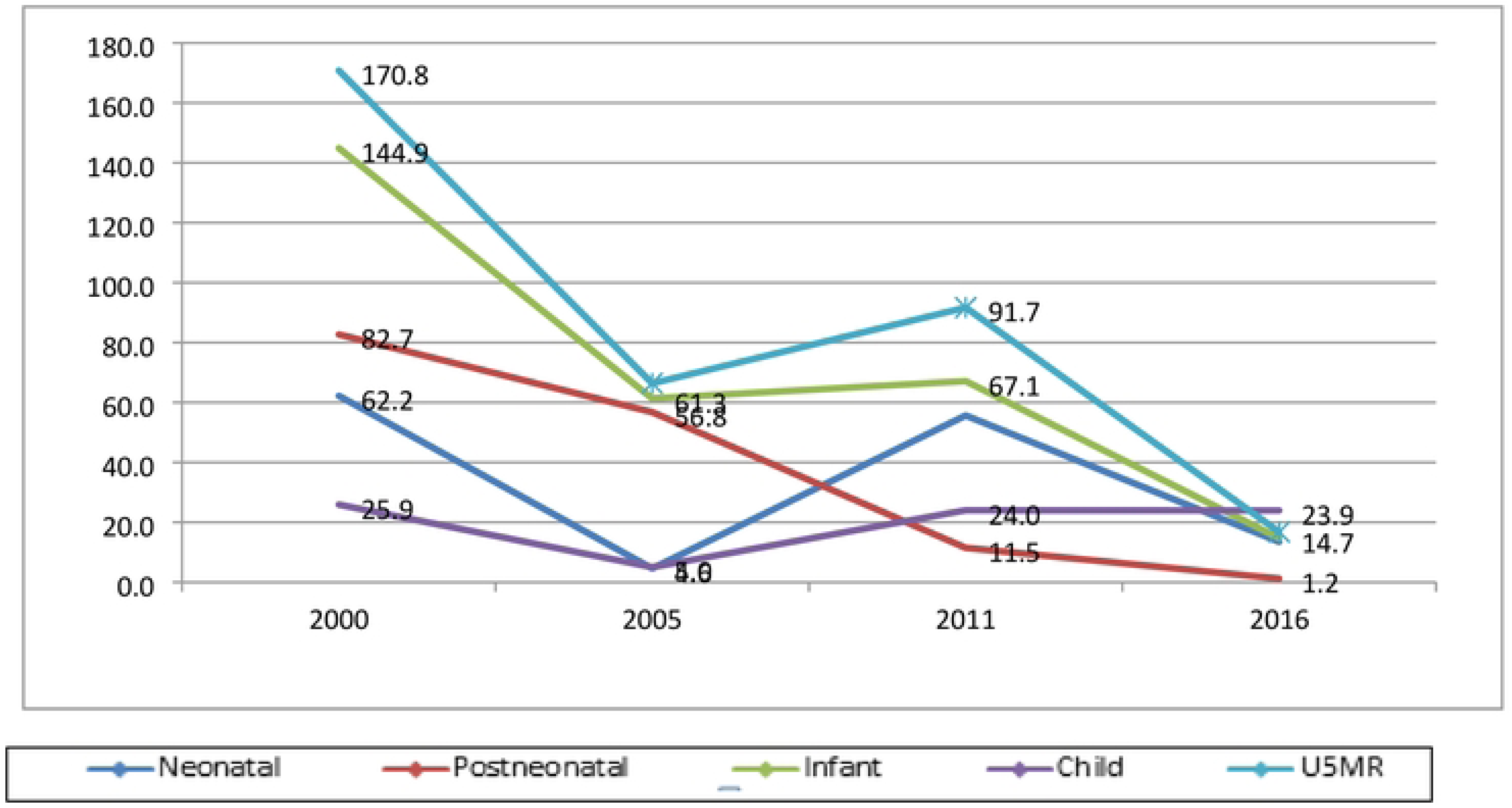
Trend of under-five, neonatal, post neonatal, infant and child mortality among children born to older women in among urban residents: Evidence from Ethiopian Demographic and Health Surveys.

Contrary to the national, urban and mainly agrarian regions the mainly pastoral region does not shows substantial decrement in absolute terms in under-five mortalities while neonatal and infant shown increment, at least in absolute terms. Accordingly, The overall mainly pastoral region under five mortality trend has shown a decreasing trend over the last 16 years from 101/1000 live births to 93 /1000 live births Infant mortality increased form 591000 live births to 81/1000 live births over the same period while neonatal mortality increased 33/1000 live births to 67/1000 live births (Fig 4).

**Fig 4:**
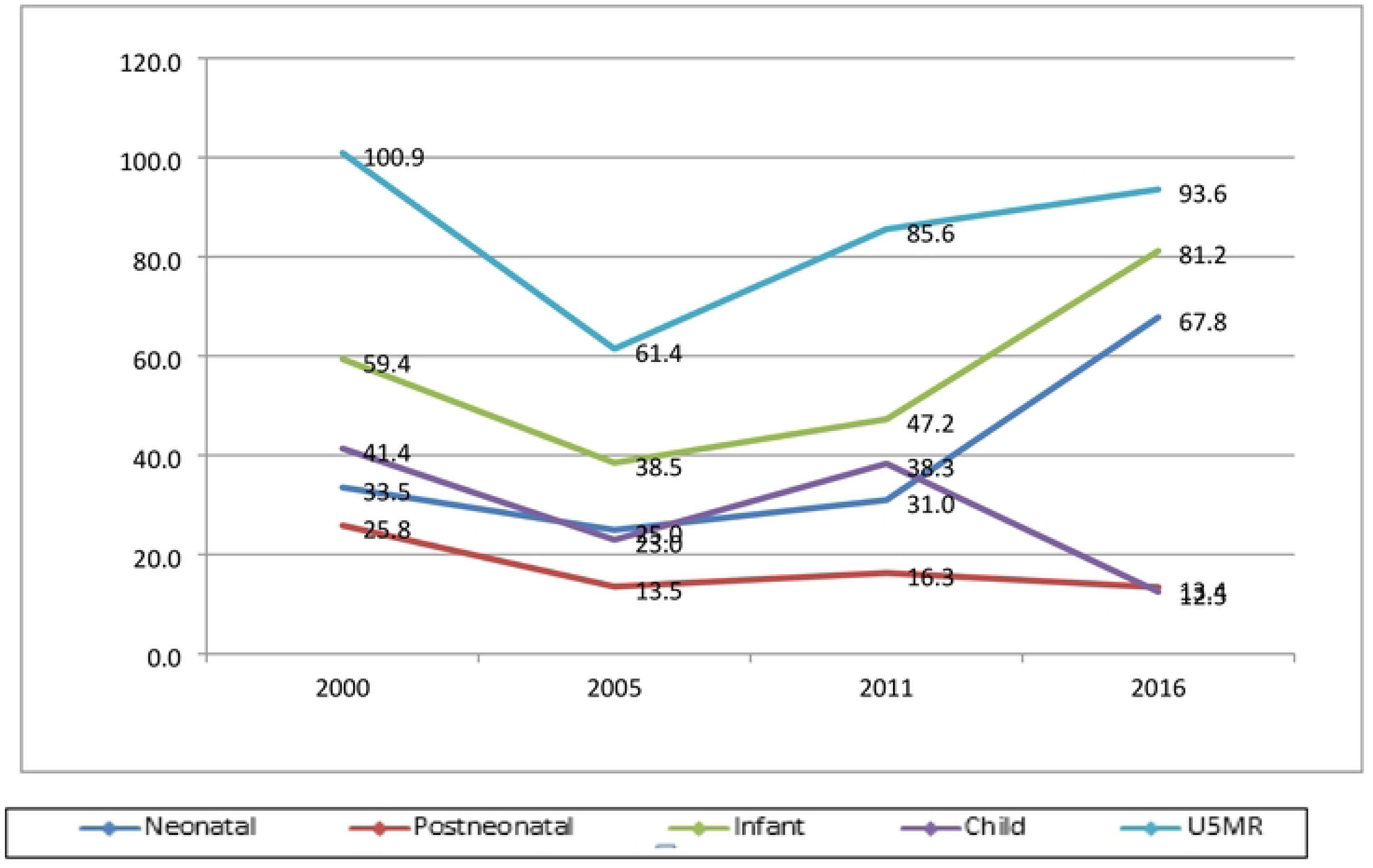
Trend of under-five, neonatal, post neonatal, infant and child mortality among children born *to* older women in Mainly Pastoralist Regions: Evidence from Ethiopian Demographic and Health Surveys.

### Multivariable Multilevel Logistics Regression

At multivariate multilevel analysis, this study identified factors contributing for under-five mortality among children of women aged 35 to 49 years old and found factors that positive and negatively statistically significant factors.

Accordingly, Sex of the child (female children), children born from first from elder mothers (31 to 40 years), children residing in family with large size (HH having a family size of 6 to 10 members), children born with longer birth interval (greater than 3 years) and children for whom their mothers received ANC visits (1 to 3 ANC vests) were found to have lower odds of mortality among children of relatively elders (Table 5).

**Table 5:**
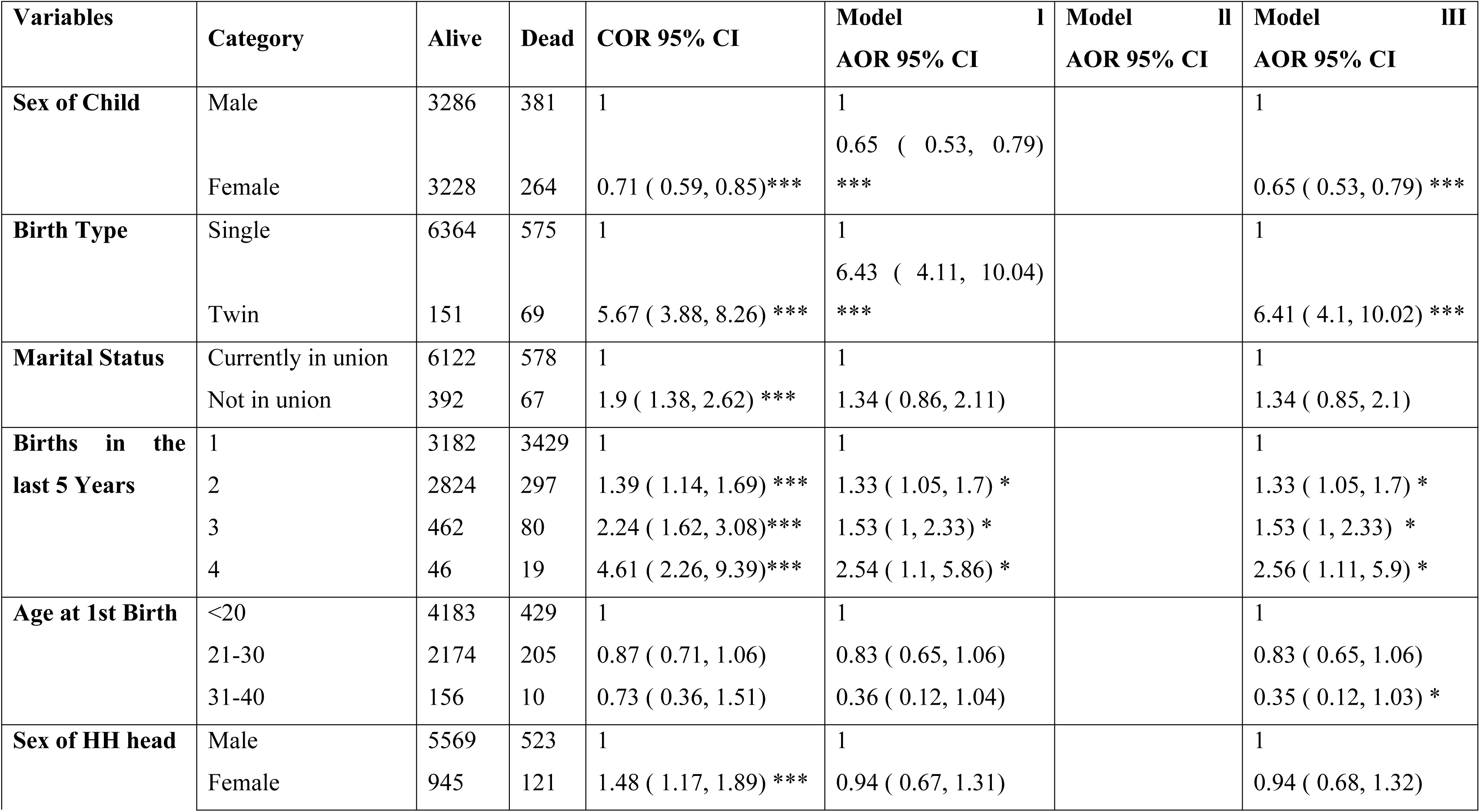
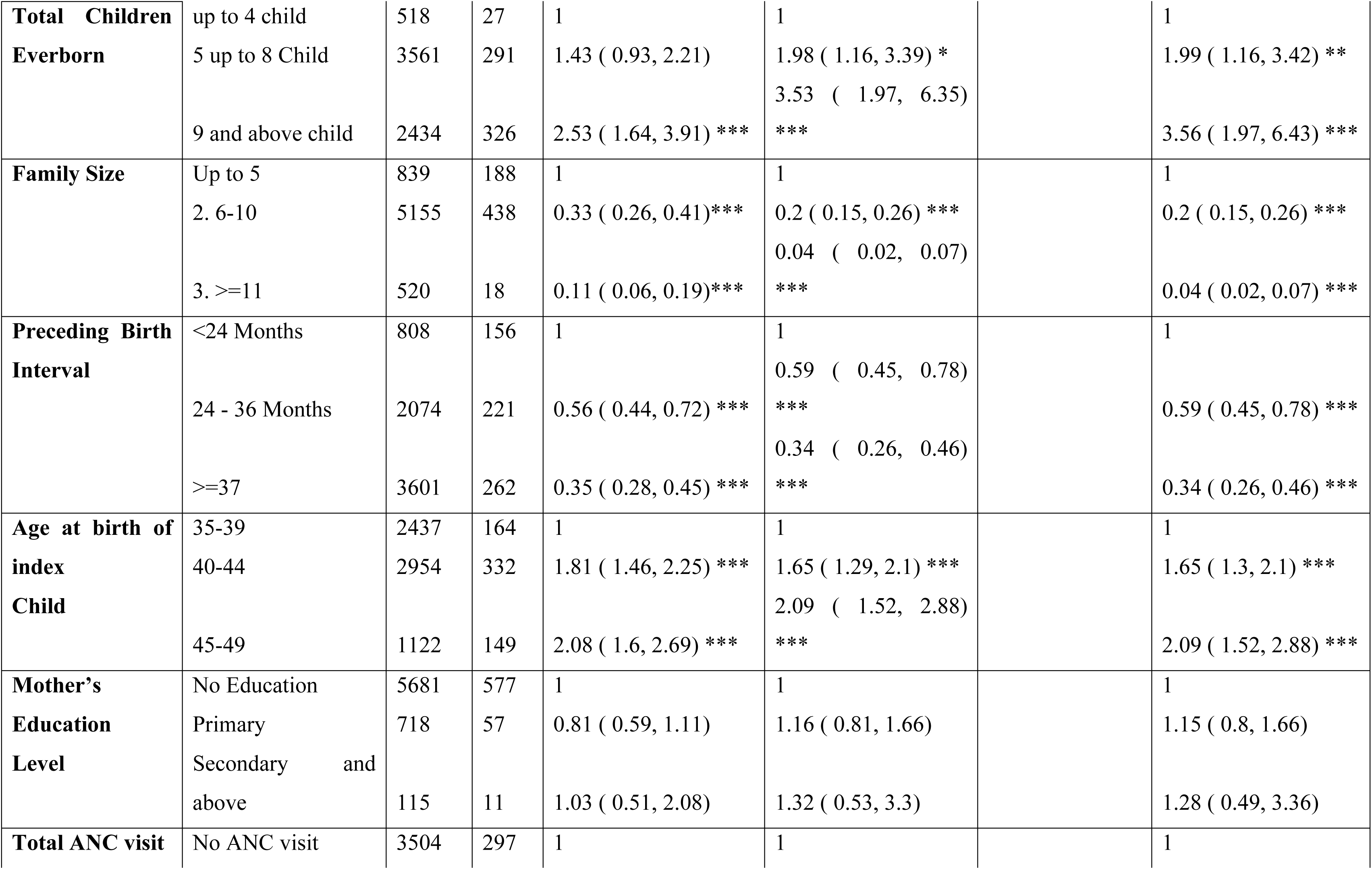
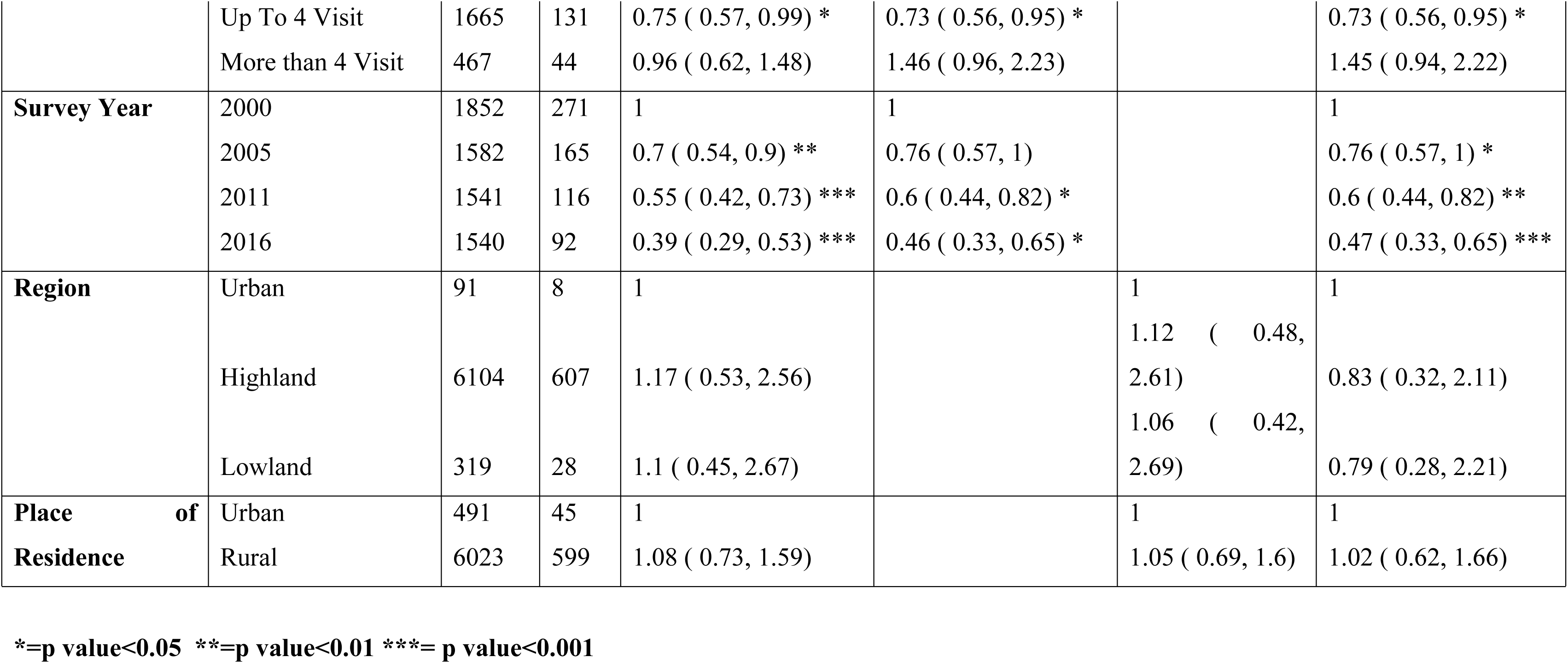
Multivariate Logistics Regression on Factors Contributing Under five mortality among children of women aged 35 to 49 In Ethiopia, Evidence from 2016 EDHS.

On the country, for women aged 35 to 49, women aged 35 to 49 years, who gave a twin birth, to those with number of births in the last five years (having 4 children), those with number of children ever born (having 9 or above children), those who gave birth of their the index child at late age (mother gave birth of the index child at late age 45 to 49) had higher odds for their children experiencing child to die before celebrating the their fifth birthday (Table 5).

The study showed that compared to male children under-five children with female gender had 35% lower AOR: 0.65 (0.53, 0.79) of being died before celebrating their 5^th^ birth day. In addition, Mothers who gave birth of their first birth at late age, 31 to 40 years at first birth had 65% lower AOR: 0.34 (0.11, 0.98) odds for their children experiencing child to die before celebrating the their fifth birthday compared with those who gave birth early. Similarly, as compared with children from households with smaller family size, this odds for children residing in household with larger family size, having a family size of 6 to 10 was reported 80% AOR: 0.2 (0.15, 0.26) lower. Moreover, children born with longer birth interval, greater than 3 years and those whose mothers received ANC visits, 1 to 3 ANC visits ANC visits for the index child had AOR: 0.35 (0.26, 0.47) and AOR:0.65 (0.5, 0.84) lowered odds experiencing mortality respectively compared with their counterparts (Table 5).

This study also identified set of factors that exacerbate under five mortality. On the contrary, women aged 35 to 49 years and who gave twin births recently had AOR: 6.15 (3.94, 9.6)) higher likelihood for their children to experiencing mortality before celebrating their 5^th^ birth day compared children of women who gave birth singleton birth. Similarly, children born from mothers who ∼gave birth of 4 children in the last four years and those born from mothers with high birth order (having 9 or above children) AOR: (2.64 (1.15, 6.06) and AOR: 3.79 (2.1, 6.84) higher odds of death respectively compared with their counter parts. Moreover, mother gave birth of the index child at late age 45 to 49 AOR: 2.13 (1.55, 2.93) had higher odds of for their children experiencing death before celebrating their fifth birthday compared with children who gave birth the index child by the age 35 to 39 Years (Table 5).

The intra cluster correlation coefficient (ICC) for the null model was 29.7% which indicates the contribution of the variation among EAs in which women reside for under five children death before celebrating their birth day while the individual characteristic difference accounted for the remaining variation. This contribution reduced to 28.8% after accounting both individual and EA level variables. Moreover, 3.1% of the variation in under-five children of women aged 35 to 49 years mortality is explained by both the individual and EA-level variables as indicate in the final model (model III where both the individual and group level variables are included). The best fitted model is model IV which has lowest AIC and higher log likelihood (Table 6).

**Table 6:** Model selection Regression on Factors Contributing Under five mortality among children of women aged 35 to 49 In Ethiopia, Evidence from 2016 EDHS.

|  | **Null** | **Individual** | **Group** | **Final** |
| --- | --- | --- | --- | --- |
| **ICC** | 0.2973 | 0.2886 | 0.29747 | 0.2880 |
| **PCV** | Reference | 0.0294 | -0.0005 | 0.0314 |
| **AIC** | 4100.8520 | 3621.5720 | 4106.5770 | 3627.3680 |
| **BIC** | 4114.3160 | 3796.4050 | 4106.5770 | 3822.3740 |
| **Variance** | 1.39 | 1.33 | 1.39 | 1.33 |
| **logLikelihood** | -<br>2048.4259 | -1784.7859 | -2048.2884 | -1784.6838 |

## Discussion

Under five mortality rates is one among the mile stones that indicate a countries health services access and coverage as well as economic development. This is why the MoH of Ethiopia and the international community make it a top agenda in the national essential newborn care service, community based management of childhood illness polices and strategies as well as the MDGs targets and the current SDG running till 2030. The save motherhood initiative, the Almata Primary health care declaration and all other health programs and polices make this target the center. Similarly, under five mortality is the core the research direction of the country, to mention the national EDHs surveys and national projects such as PMA as well the core indicator of passive surveillance under the routine HMIS/DHIS2. To this evidences also showed that the deaths more pronounced among children born elder women.

In midst of such high level government commitment and half in the SDG era measuring the level of under-five mortality and identifying its correlates among children born from mothers aged 35 to 49 years will provide actionable evidence for the ministry and relevant actors. Hence, this study reported the level of under-five mortality and its correlates among children elders mother aged 35 to 49 years using the recent full scale EDHS data.

This decrement in under-five mortality over the last one a half decade was in line with (23) and might be result the success of accomplishment of MDG goals while rigorous activities being implemented to sustain in the SDG period (3), the Ethiopian government commitment to reduce by articulating, implementing, monitoring and evaluation strategies and polies essential new born care and Integrated management of child hood illness (10, 24) along with the success of HSTP II (25). It can be also resulted from maternal and new born health service access, provision of delivery free of charge to mention few (15, 25), increased and access to contraceptive commodities to space pregnancies, the effective ness of the health extension program at community level care the expansion health facilities notably primary health care unit (14, 15) in terms of increase in primary hospitals along with building capacities of existing facilities such NICU department (2, 13) expansion of emergency and obstetrics surgery programs, specialty nursing programs such neonatal nurse (26) and the ministry of health effort to increase quality of care (27, 28).

The finding that the odds of under-five mortality lowered with year of survey can be attribute to the commitment of the Ethiopia Government to achieve the success of millennium development goals in the SDG era (3) as well as the government’s commitment to increase quality and primary health care services expansion as well as the articulation and implementation of essential new born and integrated management of childhood illness polices and strategies (10).

The finding that higher birth order increase odds of under-five mortality was In line with (29, 30) Higher birth order increased odds in line (22) While a study based on Ghana show that higher birth order lowered (31) unlike our study the finding form a study (20) that higher birth order lowers odds of under-five mortality. The discrepancy between factors might related with the socioeconomic status and health coverage between Ethiopian and Ghana, this might related with sociocultural difference between countries and related with male dominance in fertility (21).

The finding that the odds of under-five mortality increased with the number of birth 5 years preceding the survey was not in line (31) a study based on Ghana demographic survey show that number of birth in the last five years lowered the odds of under-five motility

The finding that giving twin birth increased the odds of under-five mortality in line with (19, 29), the While a study based on Ghana show twin birth lowered (31) while studies (20, 30) reported singleton birth has lower odds of under-five motility. The difference in study setting between Ethiopia and Ghana might explain it.

The finding 1 to ANC visits lower the odds of under-five mortality in this study while a study (22, 32, 33) reported that receiving the recommended increase the odds of child mortality before celebrating their fifth year birth day. Regardless of the discrepancy in the finding this finding is in line with the WHO recommendation on ANC and continuum of care resulted better pregnancy and delivery outcome.

Wider birth interval lowered the odds in line with (19, 33) while finding from (19) reported that longer birth interval increased odds while a study (30) reported that shorter birth interval increases odds of under-five mortality

Large family size increased the odds of under-five mortality as reported in a study (19, 20, 29) increase odds of under-five mortality unlike our study finding which reported large family size lowers the odds of under-five mortality

Being female child lowered the odds of under-five mortality is in line with (33) While a study based on Ghana show that female children had increased (31) being female increase odds in line with (22) unlike ours this studies (22, 29) reported male children are at increased while(19) reported not significant a study (30) reported that sex of the child has no effect on the odds of under-five mortality.

Unlike the finding from a study (19) breast feeding, source of drinking water, and income of mother were not found significant in this study. The discrepancy might be related with the differences

education and residence were not found to either negatively or positively influence death under of children before celebrating their 5th birth day which is in line (19, 22) while a study reported (30) children born from women with no education had high odds of death before their fifth birth day

For delivery modality via CS not in line with (22) which lower under five mortality While in line with (22) contraceptive use, private facility delivery and marital status increase odds of mortality unlike in our study.

Unlike our study this study (29, 34, 35) reported that rural residence and not breast feeding increase of odds of under-five death. unlike finding from our study the finding a study (20) reported that children from urban communities and severed from protected water source had lower odds of dying before celebrating their fifth birth day. While this same study reported that children were born at home had higher odds of delivery unlike our study.

Unlike ours finding a study (30) reported that that age at first birth and region of residence increase the odds of under-five mortality.

## Conclusions

The absolute number of under five deaths is 128/1000 live births in the year 2000 which reduce to 56/1000 live births in 2016. Female children, children born from first from elder mothers, those children residing am family with large size, children born with longer birth interval and children for whom their mothers received ANC visits were found to have lower odds of mortality among children.,

Women who gave a twin birth, to those with 4 births in the last five years, those with higher birth order, those who gave birth of their the index child at late age had higher odds for their children experiencing child to die before celebrating the their fifth birthday. The finding that a little higher than 1 in 11 children died before celebrating their fifth birthday in Ethiopian calls the MOH to strengthen essential new born care services, integrated management of childhood illness, increased skilled delivery to mention few.

Activities and efforts that increase uptake, promoting spaced and intended, pregnancies, advising women the better reproductive timeline to have babies, planning their family size are hoped to combat this huge surge in under five morality. Advise on pregnancy status such as type of pregnancy so that women will be aware and take necessary precaution including a must to seek skilled and facility delivery.

## Data Availability

The datasets generated during the study are publicly available from Measure DHS website. https://dhsprogram.com/data/available-datasets.com

https://dhsprogram.com/data/available-datasets.cfm

## Declarations

### Consent for publication

N/A not applicable

### Availability of data and materials

The datasets generated during the study are publicly available from Measure DHS website. https://dhsprogram.com/data/available-datasets.cfm

### Author Contributions

**Conceptualization**: Tamerat Denekew Temesegen and Tariku Dejene Demissie

**Data curation**: Tamerat Denekew Temesegen and Tariku Dejene Demissie

**Formal analysis**: Tamerat Denekew Temesegen

**Investigation**: Tamerat Denekew Temesegen

**Methodology**: Tamerat Denekew Temesegen

**Software**: Tamerat Denekew Temesegen, Solomon Abrha Damtew, Tariku Dejene Demissie.

**Supervision**: Solomon Abrha Damtew, Tariku Dejene Demissie

**Resources:** Tamirate Denekew Temesegen

**Project administration**: Tamirate Denekew, Solomon Abrha Damtew,

**Validation**: Tariku Dejene Demissie, Solomon Abrha Damtew

**Visualization**: Tamirate Denekew Temesegen, Solomon Abrha Damtew, Tariku Dejene Demissie.

**Writing – original draft:** Tamirate Denekew, Solomon Abrha Damtew, Tariku Dejene Demissie

**Writing – review & editing**: Tamirate Denekew, Solomon Abrha Damtew, Tariku Dejene Demissie.

All authors have reviewed and approved the final version of the manuscript.

## Acknowledgement

We acknowledge the Measure DHS project for providing the data.

## List if Acronyms and Abbreviations

AAU: Addis Ababa University
ANC: Antenatal care
CoDS: College of Developmental Studies
COVID: Coronavirus Disease
CSA: Central Statistical Agency
CM: Child mortality
CPR: Contraceptive Prevalence Rate
DHS: Demographic and Health Survey
EA: Enumeration Area
EDHS: Ethiopian Demographic and Health Survey
EPHI: Ethiopian Pubic Health Institute
EPHC: Ethiopian Population and Housing Census
GTP: Growth Transformation Plan
HBM: Health Belief Model
HH: House Hold
IM: Infant mortality
MDG: Millennium Development Goal
NGO: Non-Governmental Organization
NM: Neonatal mortality
NMR: Neonatal mortality Rate
PNM: Post Neonatal mortality
SDG: Sustainable Development Goal
SSA: Sub-Saharan Africa
SNNPR: Southern Nation and Nationalities Region
UN: United Nation
UNICEF: United Nations International Children’s Emergency Fund
U5M: Under-five mortality
WHO: World Health Organization

## References

1. Alkema L, New JR. Progress toward global reduction in under-five mortality: a bootstrap analysis of uncertainty in Millennium Development Goal 4 estimates. PLoS Med. 2012;9(12):e1001355.

2. FMoH. RH Strategic Plan - Ethiopia 2021-2025. 2021.

3. UN. Transforming Our World: The 2030 Agenda for Sustainable Development. New York; 2015.

4. UN. The Sustainable Development Goals Report Special edition, 2023 report. 2023.

5. Central Statistical Agency Addis Ababa E, ICF TDP, Rockville M, USA,. Demographic and Health Survey 2016. 2017.

6. Central Statistical Agency. Ethiopia: 2016 Demographic and Health Survey: Key Findings. 2016.

7. Central Statistical Agency, Macro. I. Ethiopian demographic and health survey. Final report. Calverton: ICF Macro; 2005. 2005.

8. Central Statistical Servives. Ethiopia Demographic and Health Survey 2000, entral Statistical Authority Addis Ababa, Ethiopia ORC Macro Calverton, Maryland, USA May 2001. 2000.

9. Central Statistical Agebcy, ICF Macro. EthiopiaDemographic and Health Survey 2011 Central Statistical Agency Addis Ababa, Ethiopia ICF International Calverton, Maryland, USA. 2011.

10. Miller NP, Amouzou A, Tafesse M, Hazel E, Legesse H, Degefie T, et al. Integrated community case management of childhood illness in Ethiopia: implementation strength and quality of care. Am J Trop Med Hyg. 2014;91(2):424–34.

11. MoH. Community Based Newborn Care Implementation Plan. 2012.

12. WHO, UNOCEF. HANDBOOK IMCI Integrated Management of Childhood Illness. 2005.

13. FMoH. Ethiopia National Reproductive Health Strategy 2016-2020. 2016.

14. FMoH, USID, JSI. ETHIOPIA’S PRIMARY HEALTH CARE REFORM: PRACTICE, LESSONS, AND RECOMMENDATIONS.

15. Alebachew A, Abdella E, Abera S, Dessie E, Mesele T, Mitiku W, et al. Costs and resource needs for primary health care in Ethiopia: evidence to inform planning and budgeting for universal health coverage. Front Public Health. 2023;11:1242314.

16. FEDERAL NEGARIT GAZETA OF THE FEDERAL DEMOCRATIC REPUBLIC OF ETHIOPIA. Social Health Insurance Proclamation Page 5494 Proclamation No. 690/2010 2010.

17. Merga BT, Balis B, Bekele H, Fekadu G. Health insurance coverage in Ethiopia: financial protection in the Era of sustainable cevelopment goals (SDGs). Health Econ Rev. 2022;12(1):43.

18. Ali EE. Health Care Financing in Ethiopia: Implications on Access to Essential Medicines. Value in Health Regional Issues. 2014;4:37–40.

19. Ayele BA, Abebaw Tiruneh S, Azanaw MM, Shimels Hailemeskel H, Akalu Y, Ayele AA. Determinants of under-five mortality in Ethiopia using the recent 2019 Ethiopian demographic and health survey data: nested shared frailty survival analysis. Arch Public Health. 2022;80(1):137.

20. Berelie Y, Yismaw L, Tesfa E, Alene M. Risk factors for under-five mortality in Ethiopia: Evidence from the 2016 Ethiopian Demographic and Health Survey. South African Journal of Child Health. 2019;13(3).

21. Gebretsadik S, Gabreyohannes E. Determinants of Under-Five Mortality in High Mortality Regions of Ethiopia: An Analysis of the 2011 Ethiopia Demographic and Health Survey Data. International Journal of Population Research. 2016;2016:1–7.

22. Girma M, Eshete H, Asrat R, Gebremichael M, Getahun D, Awoke T. Socio-demographic and environmental determinants of under-five child mortality in Ethiopia: using Ethiopian demographic and Health 2019 survey. BMC Pediatr. 2023;23(1):294.

23. Dheresa M, Roba HS, Daraje G, Abebe M, Tura AK, Yadeta TA, et al. Uncertainties in the path to 2030: Increasing trends of under-five mortality in the aftermath of Millennium Development Goal in Eastern Ethiopia. J Glob Health. 2022;12:04010.

24. Oringanje C, Meremikwu MM, Eko H EE, Meremikwu A, Ehiri JE. Interventions for preventing unintended pregnancies among adolescents (Review) Oringanje C, Meremikwu MM, Eko H, Esu E, Meremikwu A, Ehiri JE. The Cochrane Collaboration Published by John Wiley & Sons, Ltd. 2010.

25. FMoH. Health Sector Transformation Plan II (HSTP-II). 2021.

26. Dereje Bayissa Demissie. Paradigm shifting in postgraduate nursing education in Ethiopia: a move from classroom-based lecture to hospital-based evidencebased learning by doing at St. Paul’s Hospital Millennium Medical College. 2023.

27. FMoH. Ethiopian Health Care Quality Bulletin; Continuous Health Care Quality Improvement through Knowledge Management. 2019.

28. Premji SS, Spence K, Kenner C. Call for neonatal nursing specialization in developing countries. MCN Am J Matern Child Nurs. 2013;38(6):336–42; quiz 43–4.

29. Zewudie AT, Gelagay AA, Enyew EF. Determinants of Under-Five Child Mortality in Ethiopia: Analysis Using Ethiopian Demographic Health Survey, 2016. Int J Pediatr. 2020;2020:7471545.

30. Woldeamanuel BT. Socioeconomic, Demographic, and Environmental Determinants of Under-5 Mortality in Ethiopia: Evidence from Ethiopian Demographic and Health Survey, 2016. Child Development Research. 2019;2019:1–15.

31. Aheto JMK. Predictive model and determinants of under-five child mortality: evidence from the 2014 Ghana demographic and health survey. BMC Public Health. 2019;19(1):64.

32. Gutema GD, Geremew A, Megistu DA, Dammu YM, Bayu K. Trends and Associated Factors of Under-five Mortality Based on 2008-2016 Data in Kersa Health and Demographic Surveillance Site, Eastern Ethiopia. Inquiry. 2022;59:469580221090394.

33. Bitew FH, Nyarko SH, Potter L, Sparks CS. Machine learning approach for predicting under-five mortality determinants in Ethiopia: evidence from the 2016 Ethiopian Demographic and Health Survey. Genus. 2020;76(1).

34. Yemane GD. The factors associated with under-five mortality in Ethiopia. Ann Med Surg (Lond). 2022;79:104063.

35. Wolde KS, Bacha RH. Trend and correlates of under-5 mortality in Ethiopia: A multilevel model comparison of 2000-2016 EDHS data. SAGE Open Med. 2022;10:20503121221100608.

